# Targeted screening for lung cancer with autoantibodies

**DOI:** 10.1101/2021.08.17.21262105

**Authors:** Frank M. Sullivan, Frances S. Mair, William Anderson, Pauline Armory, Andrew Briggs, Cindy Chew, Alistair Dorward, John Haughney, Fiona Hogarth, Denise Kendrick, Roberta Littleford, Alex McConnachie, Colin McCowan, Nicola McMeekin, Manish Patel, Petra Rauchhaus, Lewis Ritchie, Chris Robertson, John Robertson, Jose Robles-Zurita, Joseph Sarvesvaran, Herbert Sewell, Michael Sproule, Thomas Taylor, Shaun Treweek, Kavita Vedhara, Stuart Schembri, on behalf of the Early Diagnosis of Lung Cancer Scotland (ECLS) Team

## Abstract

Earlier detection of lung cancer is possible, but difficult and costly to achieve. Screening with Low Dose Computed Tomography (LDCT)scanning has been shown to reduce mortality by 20-25% over the past decade but uptake amongst those most likely to suffer the disease has been slow. Resource constraints and a high false positive rate have also limited adoption of LDCT in many health systems. Targeted screening of people most likely to benefit using a range of biomarkers may be one way to improve the yield and reduce the resource requirements of LDCT. Autoantibodies, which amplify the signal produced by cancer derived proteins, are present in the blood of people mounting an immune response to cancer are a potential way to select those at highest risk. We have followed up 12 208 people enrolled in the ECLS trial for three years and shown that the specificity for early stage (I &II) disease is 90.3% throughout that period. More cancers were detected in the control than the intervention arm of the trial (101V 83). Sensitivity was 77.8% after 6 months and dropped to 46.4% after 3 years. At the end of three years the hazard ratios (95%CI) for All Cause, Cancer Specific and Lung Cancer Mortality was 0.82(0.67-1.01), 0.72(0.54-0.97) and 0.70(0.46-1.08) respectively for those randomised to Early CDT testing. As a range of treatment modalities become increasingly more effective it is even more important to target LDCT on those most likely to have early stage disease. Autoantibody testing may be one method of targeting early detection on those most likely to benefit.

## Main

Although therapeutic nihilism is no longer justified since treatment has improved, lung cancer remains the most commonly diagnosed cancer and the leading cause of cancer death worldwide. ^1^,^2^ Late presentation with symptoms is the most common means of detecting the disease which reduces the likelihood of five year survival to 3% in stage IV compared to 57% in stage I disease.^3 4^ Various means of making earlier diagnoses have been tried from patient and clinician education to screening using a range of modalities.^5^ The most effective method of screening available at present is Low Dose Computed Tomography (LDCT) scanning which has been shown in several studies to reduce mortality by 20-25% in current and former smokers aged 50 to 74 years.^6,7^ The US Preventive Task Force (USPTF) recommends that screening high-risk persons with LDCT can reduce lung cancer mortality but also notes that it causes false-positive results leading to overdiagnosis and distress.^8^ A recent estimate of uptake of the recommendation by eligible patients in the U.S. was 16%.^9^ A European position statement on lung cancer screening in 2017 recommended that implementation of low-dose CT screening should start throughout Europe as soon as possible, yet no country has initiated a national screening program.^10^ Although lung cancer is more common in people living in areas of economic deprivation, responses from this group in the population to being sent a validated questionnaire e.g. the Prostate, Lung, Colorectal, and Ovarian Cancer Screening Trial Model 2012 (PCLOm2012) is generally poor though may be improved by targeted invitation materials. ^11^ Various outreach methods have been tried to lower the barriers to accessing LDCT for eligible study subjects such as CT scanning from lorries in supermarket car parks in areas of high incidence. ^12^ An alternative would be a method which was more accessible and acceptable to more people in the target group. ^13 14^ Targeting risk using phenotypic data in electronic medical records and polygenic risk models are options being explored. ^15 16^ Biomarkers may be suitable but none of the candidates have been shown to be effective so far.^17^, ^18^

The detection of Tumor Associated autoantibodies in blood is an approach that, like Faecal Immunochemical testing compared to Faecal Occult Blood testing, may more accessible and acceptable to heavy smokers in areas of socioeconomic deprivation. ^19^These proteins are produced early in tumorigenesis, being measurable up to 5 years before the development of clinical symptoms. ^20^They represent biologically amplified markers, increasing the detectable signal for the corresponding level of antigen detected and they persist in the circulation with half-lives of typically up to 30 days (figure 1a). ^21^ The EarlyCDT-Lung Test is an enzyme-linked immunosorbent assay (ELISA) that measures seven autoantibodies, each with individual specificity for the following tumour associated antigens (TAA): p53, NY-ESO-1, CAGE, GBU4-5, HuD, MAGE A4 and SOX2. ^22^ A sample is positive if at least one AAb is elevated above a predetermined cutoff. The test has been developed throughout the pre-clinical, clinical assay validation and retrospective biomarker development pathway stages. ^23^ In cohort studies it demonstrated a specificity of 91% and sensitivity of 41%. ^24^ In the two year analysis of the Early Diagnosis of Lung Cancer Scotland (ECLS) trial the EarlyCDT-Lung test had an estimated sensitivity of 52.2% (95% CI = 30.6 to 73.2) for stage I/II disease and 18.2% (95% CI = 7.0 to35.5) for stage III/IV disease, and specificity of 90.3% (stage I/II; 95% CI = 89.6 to 91.1) and 90.2% (stage III/IV; 95% CI = 89.4 to 91.0).^25 26^

**Figure 1.**
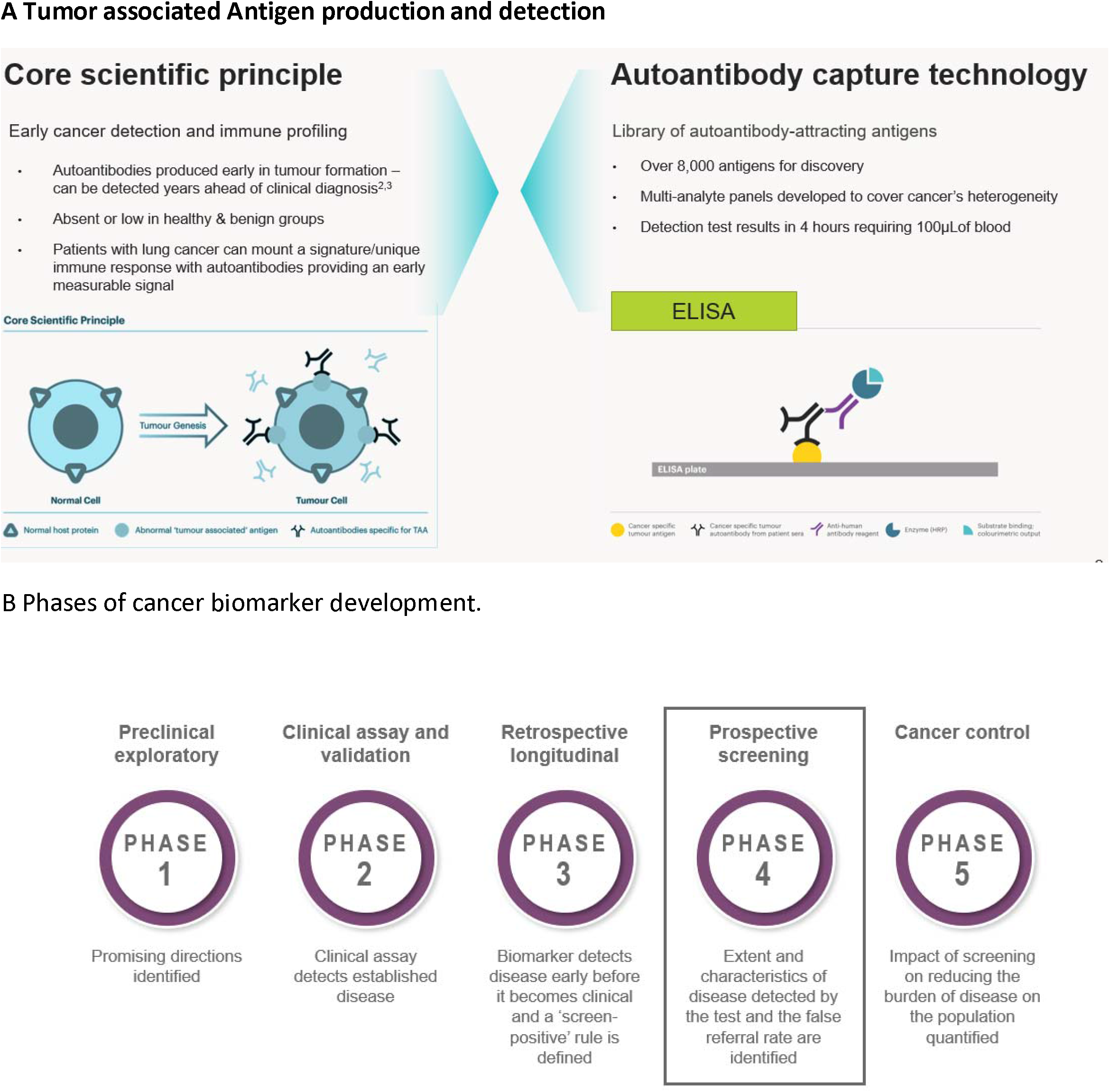
Early CDT mode of action and place of current study in the stages of biomarker development.

ECLS was a phase IV biomarker (prospective screening) study(figure 1b). ^27^ It was a randomised trial of 12 208 smokers and ex-smokers age 50–75 at risk of developing lung cancer using Early CDT followed by imaging recruited from General Practices in Scotland. The intervention arm received the EarlyCDT-Lung test and, if test positive, low-dose CT scanning six-monthly for up to two years. EarlyCDT-Lung test negative and control arm participants received standard clinical care. Outcomes were assessed at two years post-randomisation using validated data on cancer occurrence, cancer staging, mortality and comorbidities. Six monthly assessments over the same period captured participants psychological (eg mental health) and behavioural (eg smoking behaviour) outcomes. ^28,29^ After two years, there was a one third reduction in late stage cancers diagnosed (hazard ratio for stage III/IV presentation was 0.64 (95% confidence interval 0.41, 0.99)) in the intervention group compared to the controls. There were large but non-significant differences in lung cancer and all-cause mortality after two years. This paper presents data after three years of follow up on diagnosis of cancers at different stages and the effect on mortality.

## Results

### After three years, the numbers of late stage cancers and deaths were lower in patients tested for autoantibodies

Using the Scottish Cancer Registry we determined that 184 of the 12 128 study participants (1.5%) were diagnosed with lung cancer in the three years after randomisation. Table 1 shows that there were significantly more lung cancers diagnosed in the control group (101 V 83, HR2.97(1.79-4.92)) and cancer specific mortality was lower in those tested (103 V 74, HR 0.72(0.54-0.97)). Large but non-significant reductions in all cause and lung cancer deaths were also reported.

**Table 1.** Summary of participants, diagnoses and deaths in the arms of the trial.

|  | **Control**<br>Standard<br>Clinical Care | **Intervention**<br>EarlyCDT-Lung Test | **Hazard Ratio**<br>(95% CI) |
| --- | --- | --- | --- |
| Participants | 6121 | 6082 |  |
| **Diagnoses** |  |  |  |
| All Lung<br>Cancers | 101 | 83 | 2.97(1.79-4.92) |
| Stage I & II<br>lung<br>cancers | 26 | 28 | 1.08 (0.64-1.85) |
| Stage III, IV,<br>and<br>unclassified<br>cancers | 75 | 55 | 0.73(0.51-1.02) |
| **Deaths** |  |  |  |
| All cause<br>deaths | 203 | 166 | 0.82(0.67- 1.01) |
| All cancer<br>deaths | 103 | 74 | 0.72(0.54-0.97) |
| All Lung<br>cancer<br>deaths | 50 | 35 | 0.70(0.46-1.08) |

**Table 2.** Performance characteristics of Early CDT at 6 monthly intervals over the 1^st^ 3 years.

| Year since<br>randomisation | Performance<br>Characteristic | Value | Confidence Interval |  |
| --- | --- | --- | --- | --- |
|  |  |  | lower | upper |
| 0.5 | Negative Predictive Value | 0.9996 | 0.999 | 1.000 |
|  | Positive Predictive Value | 0.0117 | 0.024 | 0.005 |
|  | Sensitivity | 0.7778 | 0.400 | 0.972 |
|  | Specificity | 0.9028 | 0.910 | 0.895 |
| 1 | Negative Predictive Value | 0.9993 | 0.998 | 1.000 |
|  | Positive Predictive Value | 0.0151 | 0.028 | 0.007 |
|  | Sensitivity | 0.6923 | 0.386 | 0.909 |
|  | Specificity | 0.9030 | 0.910 | 0.895 |
| 1.5 | Negative Predictive Value | 0.9980 | 0.996 | 0.999 |
|  | Positive Predictive Value | 0.0201 | 0.035 | 0.010 |
|  | Sensitivity | 0.6875 | 0.413 | 0.890 |
|  | Specificity | 0.9033 | 0.911 | 0.896 |
| 2 | Negative Predictive Value | 0.9980 | 0.996 | 0.999 |
|  | Positive Predictive Value | 0.0201 | 0.035 | 0.010 |
|  | Sensitivity | 0.5217 | 0.306 | 0.732 |
|  | Specificity | 0.9034 | 0.911 | 0.896 |
| 2.5 | Negative Predictive Value | 0.9974 | 0.996 | 0.999 |
|  | Positive Predictive Value | 0.0201 | 0.035 | 0.010 |
|  | Sensitivity | 0.4615 | 0.266 | 0.666 |
|  | Specificity | 0.9033 | 0.911 | 0.896 |
| 3 | Negative Predictive Value | 0.9973 | 0.995 | 0.998 |
|  | Positive Predictive Value | 0.0217 | 0.037 | 0.012 |
|  | Sensitivity | 0.4643 | 0.275 | 0.661 |
|  | Specificity | 0.9035 | 0.911 | 0.896 |

### Autoantibodies detected by EarlyCDT-Lung are specific and most sensitive for early stage disease in the first year after testing

Test positives (n=598 (9.8%) of the 6088 tested) received a more intensive intervention than those who tested negative since those who were test negative were treated like the control group. The specificity for early stage (I &II) disease was 90.3% throughout that period. Estimated sensitivity for early stage disease dropped from 77.8% after 6 months to 46.4% after 3 years.

### Lung cancers detected by EarlyCDT-Lung were mainly early stage

The cumulative incidence of lung cancers (All, Early, Late) over time in the Intervention (test positive and test negative) and control groups over the 3 year follow up period is shown in figure 2 below. This shows that the main effect of the test was to diagnose a substantial proportion of the early stage cancers within the first six months days after the test. Thereafter few early cancers were diagnosed and late cancers became evident. This suggests autoantibodies detect early stage disease more effectively than late stage disease. If the test were used in a screening program it would need to be repeated at intervals to detect incident disease.

**Figure 2.**
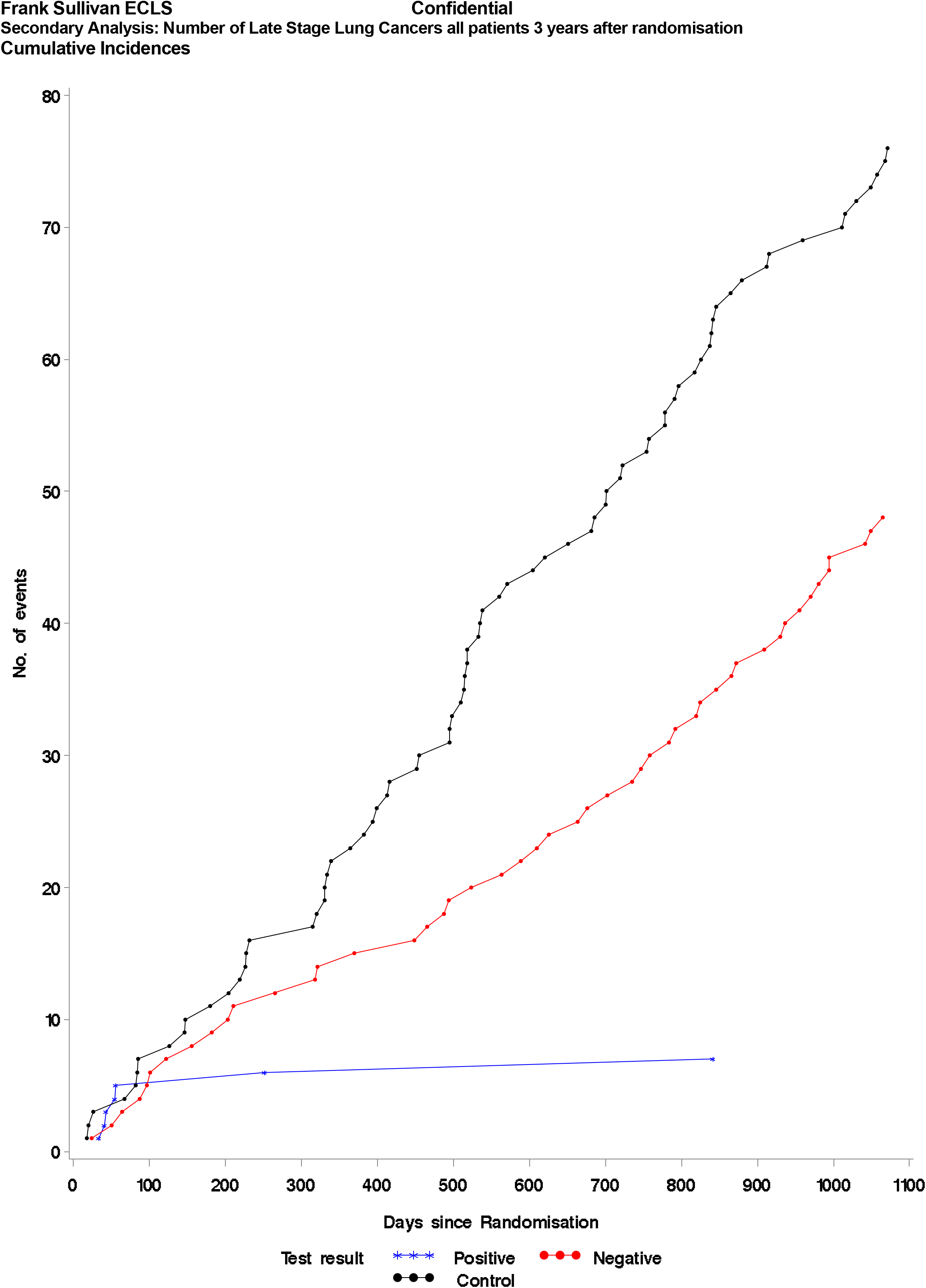
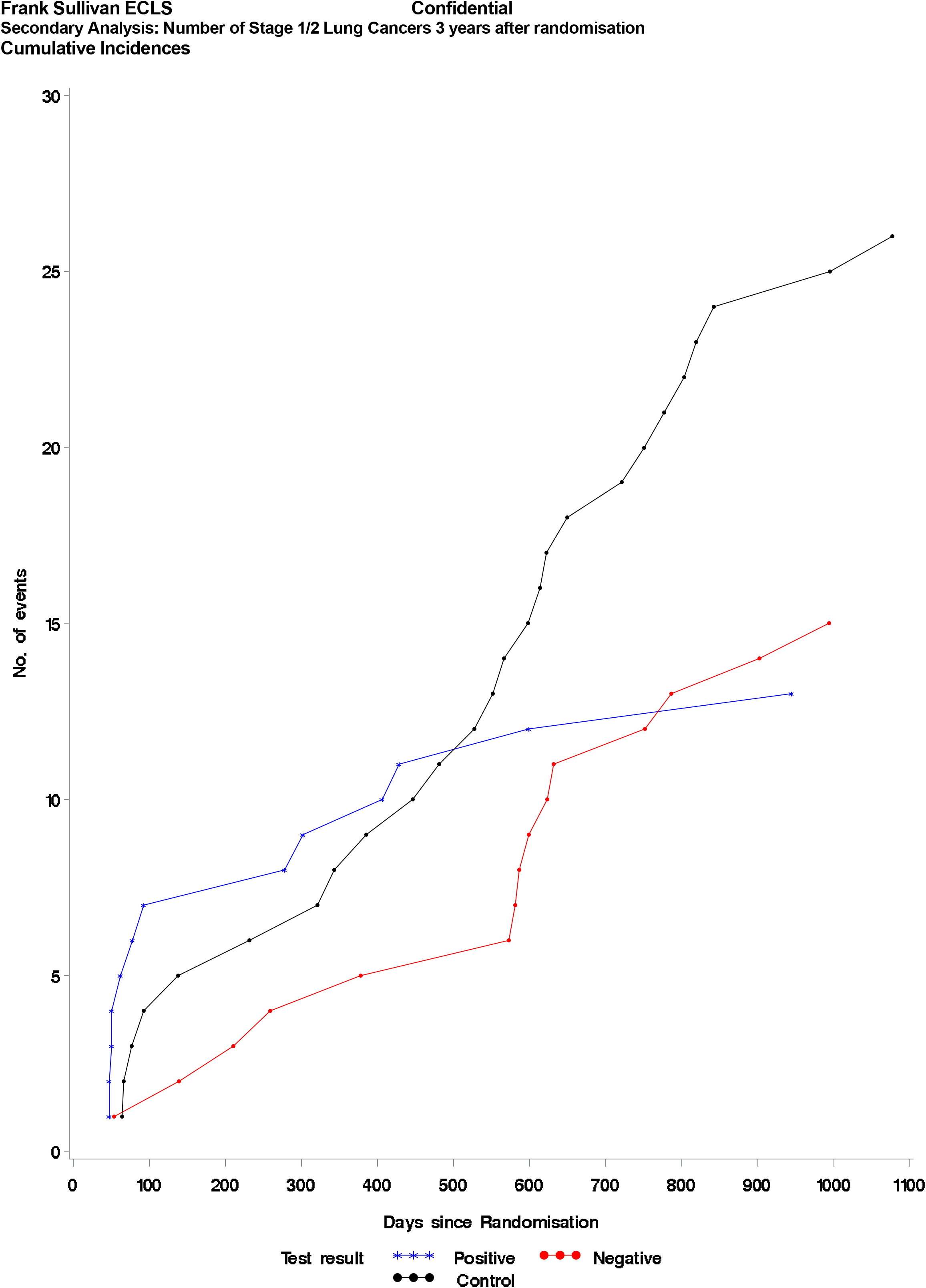
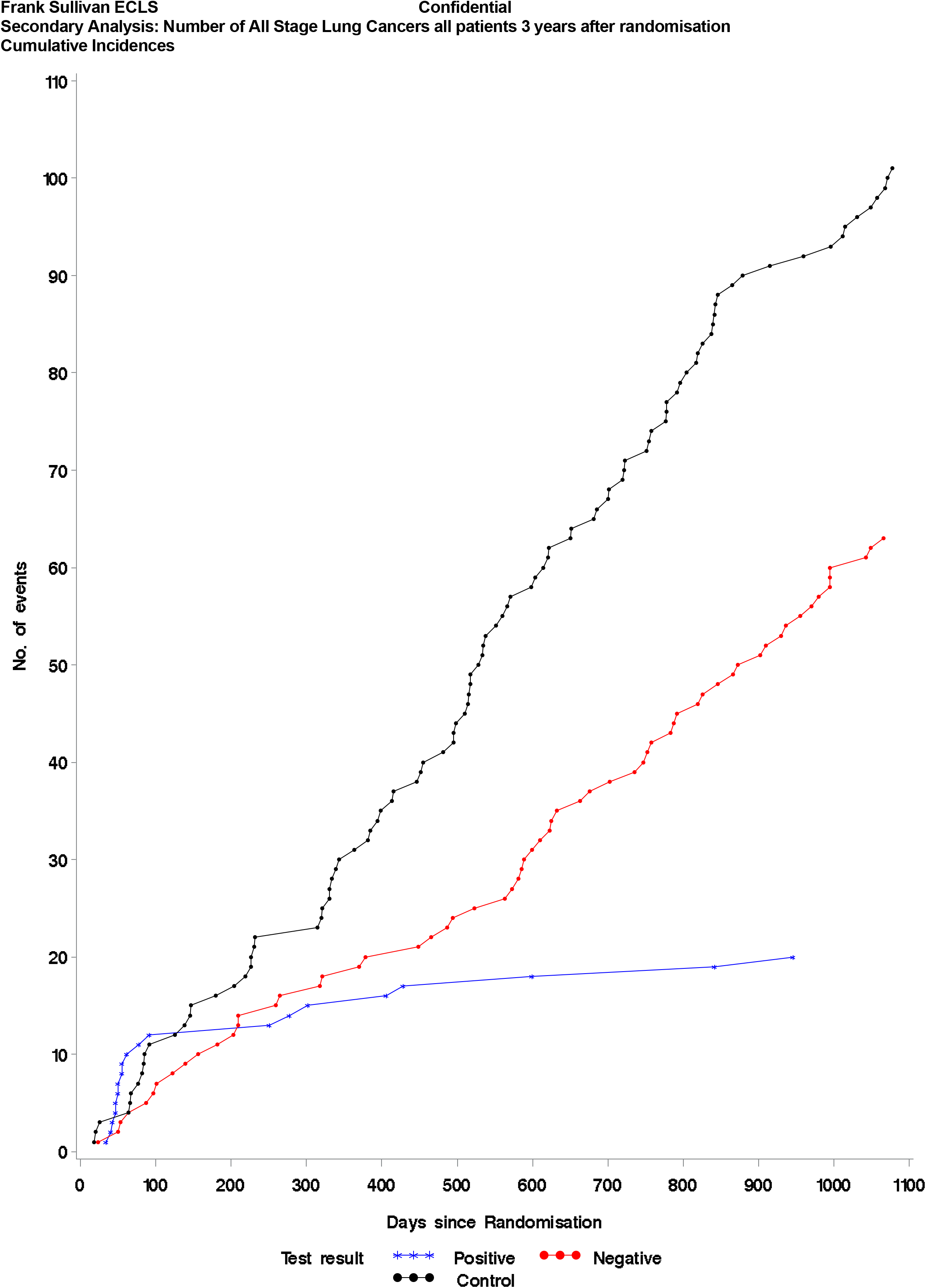
Diagnosis of lung cancer 3 Years After Randomisation in the Intervention (Test +ve and Test -ve) and Control Arms. a. All lung cancers b. Early Stage Diagnoses C. Late stage Diagnoses

### All cause, cancer specific and lung cancer mortality was reduced

The cumulative mortality (All cause, Cancer specific, Lung cancer) over time in the Intervention (test positive and test negative) and control groups over the 3 year follow up period is shown in figure 3 below. The Hazard ratio (HR) for all cause mortality comparing the intervention and control groups was 0.82().67-1.01). For cancer specific mortality the HR was 0.72(0.54-0.97) and for Lung cancer mortality it was 0.70(0.46-1.08),

**Figure 3.**
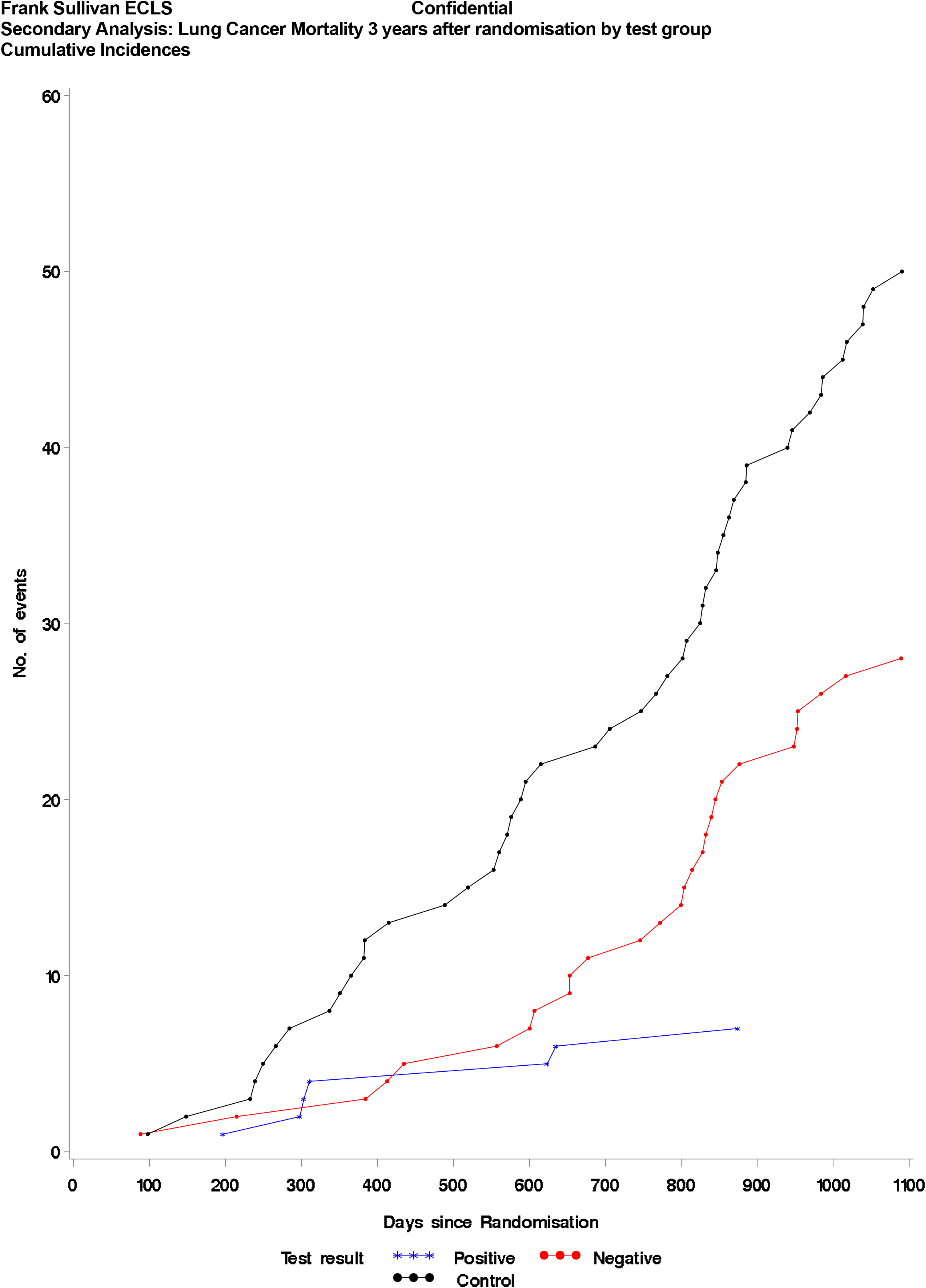
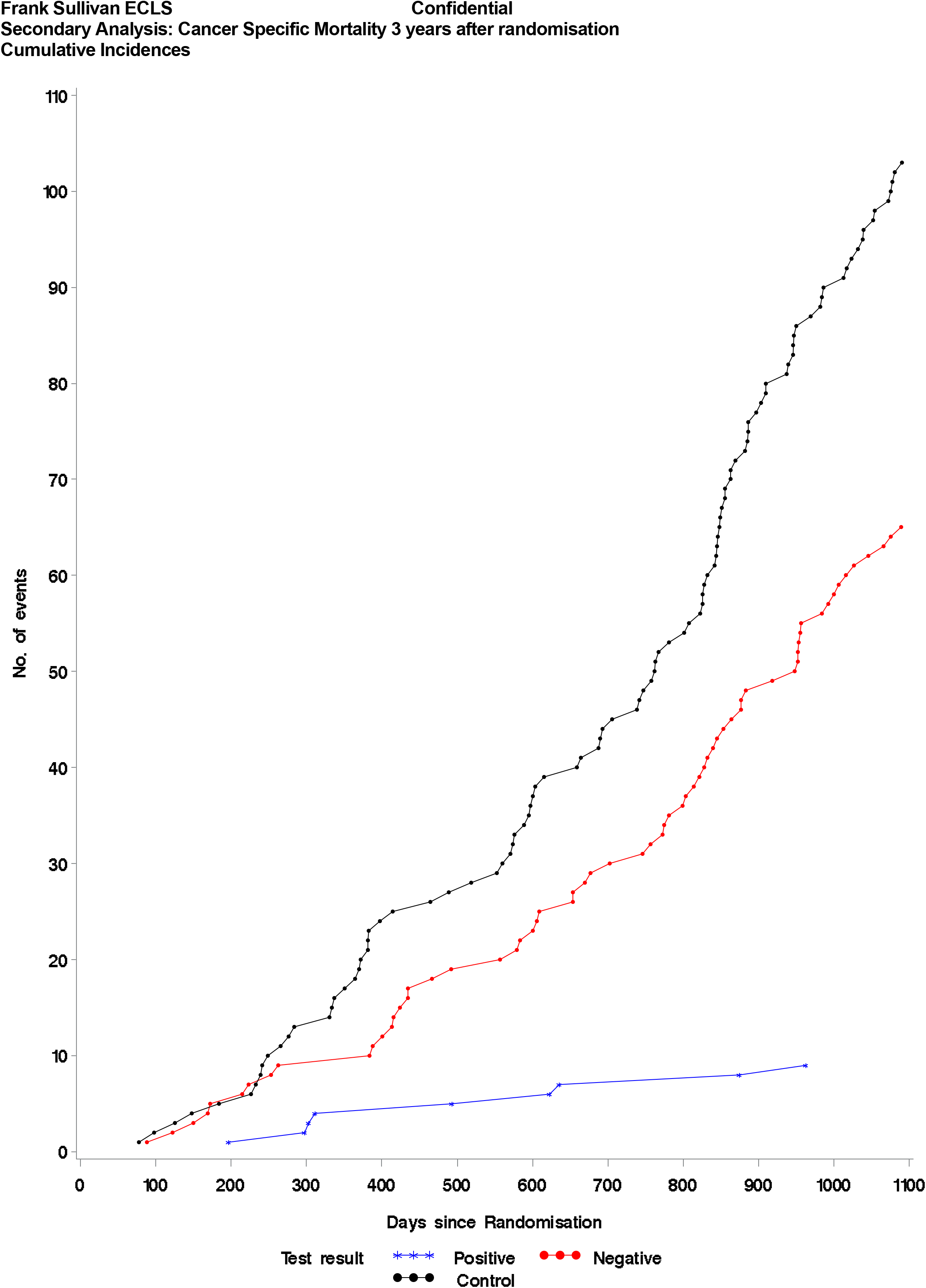
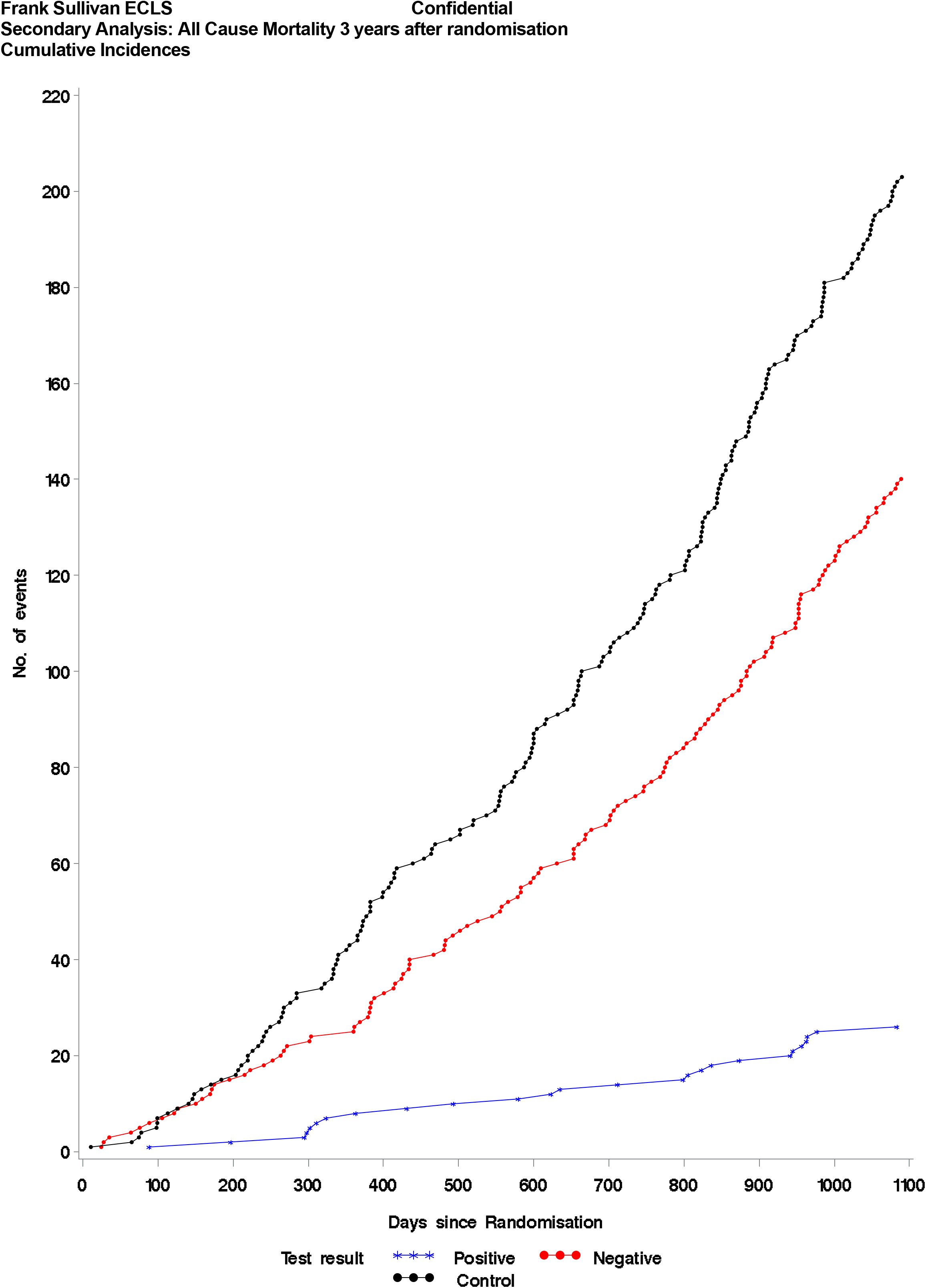
Mortality in the Intervention (test +ve and test-ve) groups and the control group. a. All Cause Mortality b. Cancer Specific Mortality c. Lung Cancer Mortality

### The number needed to screen to detect one early stage cancer is 472

The number needed to screen (NNS) to avoid one late stage cancer by Early CDT followed by LDCT compared to the control group was calculated as the reciprocal of the absolute risk reduction to be 295 (−24 to 615). Assuming the 3 year survival from lung cancer increases from 20% at present to 80% if tested this suggests a NNS to avoid 1 death of 472.

## Discussion

We have presented three year follow-up data from an RCT comparing a single autoantibody test compared to standard clinical practice over three years in a pragmatic study design. ^30^ The main findings are that after three years, the numbers of late stage cancers and deaths were lower in patients tested for autoantibodies. Autoantibodies detected by EarlyCDT-Lung are specific and most sensitive for early stage disease in the first year after testing and the cancers detected were mainly early stage. All cause, cancer specific and lung cancer mortality was reduced with a number needed to screen to detect one early stage cancer of 472.

Strengths of this study include community based recruitment with a clear denominator; a high proportion of participants recruited from the two most socioeconomically deprived quintiles (51.8%) of the Scottish population; integration within a National Health Service providing whole population care; a high end-point ascertainment rate (>99.9%); and analysis based on the groups to which patients were allocated.

The test performance of potential biomarkers should be compared ideally with the gold-standard method for the clinical application of interest but this is not always possible. ^31^No gold standard exists for the earlier diagnosis of lung cancer at present. ^32^,^33^ In our study, participants who tested negative by EarlyCDT-Lung and those in the control arm, were not offered LDCT. This could be considered a limitation of our study but offering LDCT to smokers aged 50-75 was not, and is still not, the standard of care in NHS Scotland when the study was designed and approved, nor was there capacity within the service to undertake the number of scans required. ^34^ Our follow up period of three years was relatively short and cases will continue to emerge as the study final results after ten years of follow up become available. Another limitation is that we have reported stage at diagnosis and mortality in patients diagnosed between two and three years of follow up. This will tend to reduce the apparent effectiveness of the test. The results of this study are not directly comparable to those using a validated questionnaire to identify people wo may be eligible for LDCT screening. ^35^ We are planning a direct comparison of both methods to determine how a biomarker test compares to a questionnaire followed by LDCT. Whether the optimal testing interval may be one or two years will also be tested.

## Methods

### Study Design and Dataset

The methods used in the ECLS study are described in the protocol.^36^ Validated data on cancer occurrence, mortality and comorbidities were obtained, with patient consent, from National Services Scotland, which is a high-quality health services data repository.^37^ These were linked deterministically to baseline and follow up visit data in OpenClinica using Scotland’s Community Health Index number and analysed in the Dundee Health Informatics Centre Data Safe Haven. ^38^ Pathology and tumour staging reports were prepared by independent assessors who were blinded to the allocation status of study participants. Staging data were taken from the Scottish Cancer Registry (SMR06). The primary outcome variable in the trial was the first occurrence of cancer diagnosis using the International Statistical Classification of Diseases and Related Health Problems 10th Revision codes (ICD-10) C33 (primary malignant neoplasm of trachea) and C34 (bronchus or lung). Where more than one lung cancer tumour was present at diagnosis, the most advanced tumour was used for classification of disease at diagnosis. To determine staging, reported clinical and pathological “T, N, M” were used with pathological staging taking precedence when present by data analysts blinded to allocation status. Lung tumour histology was coded in accordance with the Third Edition International Classification of Diseases for Oncology and lung cancer staging was determined using TNM 7th Edition.^39^

The analyses followed the intention to treat principle with a subgroup analyses of those who tested positive or negative in the intervention arm. Cox proportional hazards models were used to estimate the hazard ratios. One participant who withdrew consent for use of their data was excluded from analysis. The models were adjusted for age, gender, smoking history, socioeconomic status and General Practice. Comparisons of proportions were carried out using Fisher’s exact test due to the small number of events. Poisson regression models, (adjusting for follow-up time when necessary) were used to investigate other clinical outcomes. Further details are available in the Statistical Analysis Plan appendix).

Specificity and sensitivity were estimated from cancer registry (SMR06 data) which, in this prospective study, used cancer status determined at six monthly intervals. Follow-up was performed using a national, closed administrative data system for 36 months after individual randomisation or to death if within the follow-up period. We also checked national prescribing, and inpatient and outpatient data systems for activity relating to trial participants in the two-year post-randomisation follow-up period.

## Data Availability

The dataset is governed by data usage policies specified by the data controller (Tayside Academic Health Sciences Collaboration). We are committed to complying with the UK Policy Framework for Health and Social Care Research. Data from this study will be available for commercial and non-commercial research purposes upon approval by the study Data Access Committee according to institutional requirements. Applications for data access should be directed to.

https://eclsstudy.org/

## Data Availability

https://eclsstudy.org/

## Data availability

The dataset is governed by data usage policies specified by the data controller (Tayside Academic Health Sciences Collaboration). We are committed to complying with the UK Policy Framework for Health and Social Care Research. ^40^ Data from this study will be available for commercial and non-commercial research purposes upon approval by the study Data Access Committee according to institutional requirements. Applications for data access should be directed to.

## Acknowledgements

Funding has been received from the Scottish Government Health and Social Care Directorate and

Oncimmune. Funding information for this article has been deposited with the Crossref Funder Registry.

